# Intraluminal thrombus effect on the progression of abdominal aortic aneurysms by using a multistate continuous-time Markov chain model

**DOI:** 10.1101/2020.08.31.20185330

**Authors:** Liangliang Zhang, Byron A. Zambrano, Jongeun Choi, Whal Lee, Seungik Baek, Chae Young Lim

**Affiliations:** Department of Biostatistics, University of Texas MD Anderson Cancer Center; Department of Mechanical Engineering, Texas A&M University; School of Mechanical Engineering, Yonsei University; Department of Radiology, Seoul National University Hospital; Department of Mechanical Engineering, Michigan State University; Department of Statistics, Seoul National University

**Keywords:** Abdominal aortic aneurysms, Intraluminal thrombus, Markov chain model, survival analysis

## Abstract

**Objective:** Recent studies reported the intraluminal thrombus (ILT) among others strongly affects abdominal aortic aneurysms (AAA) expansion rates. Thus, we investigate characteristics of ILT with AAA expansion.

**Methods:** We applied homogeneous multistate continuous-time Markov chain models to longitudinal data of 26 Korean AAA patients as a retrospective clinical study. We considered four states of AAA and maximal thickness of ILT (max_ILT_), fraction of wall area covered by ILT (area_frac_) and fraction of ILT volume (vol_frac_) as possible covariates.

**Results:** Based on likelihood-ratio statistics, area_frac_ is the most significant biomarker and max_ILT_ is the second most significant. Besides, within AAAs that developed an ILT layer, we found that an AAA expands relatively fast at an early stage but the rate goes slower once AAA reaches in a large size.

**Conclusion:** Results recommends surgical intervention when any patient with area_frac_ more than 60% or max_ILT_ more than 30mm. Although this recommendation should be considered with caution given the limited sample size, one can use the proposed model as a tool to find such recommendations with their own data.

## Introduction

Abdominal aortic aneurysm (AAA), the dilatation of an aorta at an abdominal level is a common threatening disease that affects 9.5% of the elderly population (>65 years)^1^. This dilatation of the abdominal aorta can cause death when it ruptures. The rupture occurs when the stress on the AAA wall overcomes the wall strength. Since the maximum diameter is being positively associated with the wall stress^2^ and with AAA expansion^3^, it has been used as a risk factor for AAA growth and subsequently for rupture. Current medical treatment suggests that once a patient is diagnosed with AAA, s/he should be under surveillance until the aneurysm reaches 5.5 cm in diameter^4^ in the U.S. Some studies, however, challenge the 5.5 cm threshold criterion since small AAAs still rupture. For example, 10–24% of ruptured AAAs were less than 5 cm in diameter^5^, although more cases succumb to rupture prior to surgical intervention as the diameter increases. In fact, the screening criterion varies by countries^6^. Therefore, we take advantage of this association and investigate the statistical distribution of the maximum diameters of AAAs by quantitatively dichotomizing the patients into four different prognosis groups: early, mild, severe and fatal.

In this study, patients were observed over time and morphological characteristics of AAA were collected as well. With the time-to-event data, researchers and practitioners usually would like to make inferences on the progression path (e.g. deterioration or recovery) of the disease. Survival analysis is a commonly used statistical tool to achieve such goal. However, usual survival analysis can only deal with binary events (alive or death). Instead, the multistate model^7^ is able to model longitudinal studies where individuals may experience several events, and conduct forward or backward conversions between events. Therefore, we adopted a multistate continuous-time Markov Chain model to estimate the transition from one state to another for AAA. In this model, the transition intensities provide the hazards^8^ and the survival probabilities for AAA progression. We can also calculate the mean sojourn time^9^ in a given state. In addition, we incorporate the intraluminal thrombus (ILT) information as covariates into the model through transition intensities as ILT is strongly associated with AAA expansion rates.

Choosing the appropriate covariate to predict AAA growth rate and time-to-event is an important task, because an accurate prediction can allow personalized clinical management and proper timing of surgery^10^. Patient-specific prediction of AAAs have been performed via Bayesian methods combining sequential CT images^11^ and biomedical and computational models^12^ without taking into account the intraluminal thrombus (ILT). There has been, however, substantial heterogeneity of AAA expansion rates and an accurate growth prediction is still remaining a top priority to improve the prediction capability. Numerous variables have been suggested as potential predictors for growth and its rupture^13^ and, among them, the ILT is being recognized as a potential metric of AAA growth^14^. For examples, Stevens *et al*.^15^ obtained a set of follow-up CT images from four patients and estimated AAA growth along with geometrical characteristics of ILT. They found that the intraluminal ILT volume and maximum ILT thickness were correlated with AAA volume growth. With 14 patients, Zambrano *et al*.^16^ classified scan images of AAAs into two groups of AAAs by the ILT areal fractions and found that AAAs grew significantly faster in the group with larger ILT compared to the group with lesser ILT. Nevertheless, those studies of ILT on AAA growth were found only from on a small set of AAAs and few studied on which geometrical characteristics of ILT were more linked to the prediction of AAA expansion. Therefore, using a larger dataset of follow–up AAAs, this study calls to investigate which geometrical characteristics of ILT can enhance the prediction of AAA expansion and their transition probabilities. Particularly, we study the association between AAA growth and ILT by considering a homogeneous multistate continuous-time Markov chain model with covariates related to ILT.

Generally, the progression of AAA growth can be related to other factors such as age, gender, smoking, and vascular diseases^17^. Our retrospective data, on the other hand, were originally obtained for characterizing relevant morphological parameters so that the clinical and biochemical data were not available. As our focus is on influence of ILT information for AAA progression, we extracted ILT thicknesses, percentage of the luminal area covered during the AAA progression and the volume of ILT per each scan and applied a statistical model to predict AAA expansion based on these geometrical characteristics of ILT. By incorporating contributing factors such as ILT’s volume or fraction, a fitted model can be used to predict the progression of AAA and its rate from one state to another state.

## Materials & Methods

This study uses longitudinal computed tomography (CT) images from 26 de-identified Korean patients with each patient having up to 7 surveillance scans. Thus, this study is the retrospective review of clinical data. The study was reviewed and exempted by Internal Review Board at Michigan State University. For each scan, a maximum spherical diameter *D* was measured and used to categorize AAAs into 4 stages^18^: 1) ‘early’ (*D* < 40mm), 2) ‘mild’ (40mm ≤ *D* <47mm), ‘severe’ (47mm ≤ *D* < 58 mm), 4) ‘fatal’ (*D* ≥ 58mm). Gharahi *et al*.^18^ proposed a diameter measurement called the spherical diameter that the largest sphere fits within the aorta and found that the spherical diameter measurement has the minimum uncertainty compared to others, which has an advantage for the growth prediction, although the spherical diameters are slightly smaller than the traditional orthogonal diameters (average 4 mm with the maximum value of 9 mm). Note that there are variations in AAA screening among countries. However, the typical orthogonal diameter criterion for surgical recommendation is 50mm or 55mm^4,6^. Given such variations in criterion and use of spherical diameter in our study, it is reasonable to assume that a patient in severe state is recommended for surgical intervention and a patient in fatal state should be guided for a prompt surgery.

Also, for each scan, maximal thickness of ILT, fraction of wall area covered by ILT and fraction of ILT volume were measured by the approach introduced in Zambrano *et al*. (2016)^16^ and denoted by max_ILT_, area_frac_ and vol_frac_, respectively. AAA maximum spherical diameter information of all patients at different scan times are shown in Figure 1. There are cases where a state of AAA growth is moved to another state (e.g. move from state ‘early’ to state ‘mild’ or from state ‘mild’ to state ‘severe’.). Note that one patient (P14) experienced a decrease of the maximum diameter before an expansion. There are also cases that scan times are relatively short. One patent (P21) has much smaller diameter compared to the other patients. The measurements of this patient could be an outlier or due to the characteristics of the spherical diameter (smaller than that of the traditional orthogonal diameter^18^). As P21 stayed in the “early” state according to our criterion, the results would not change much with/without this patient so we decide to keep the patient in the analysis.

**Figure 1.**
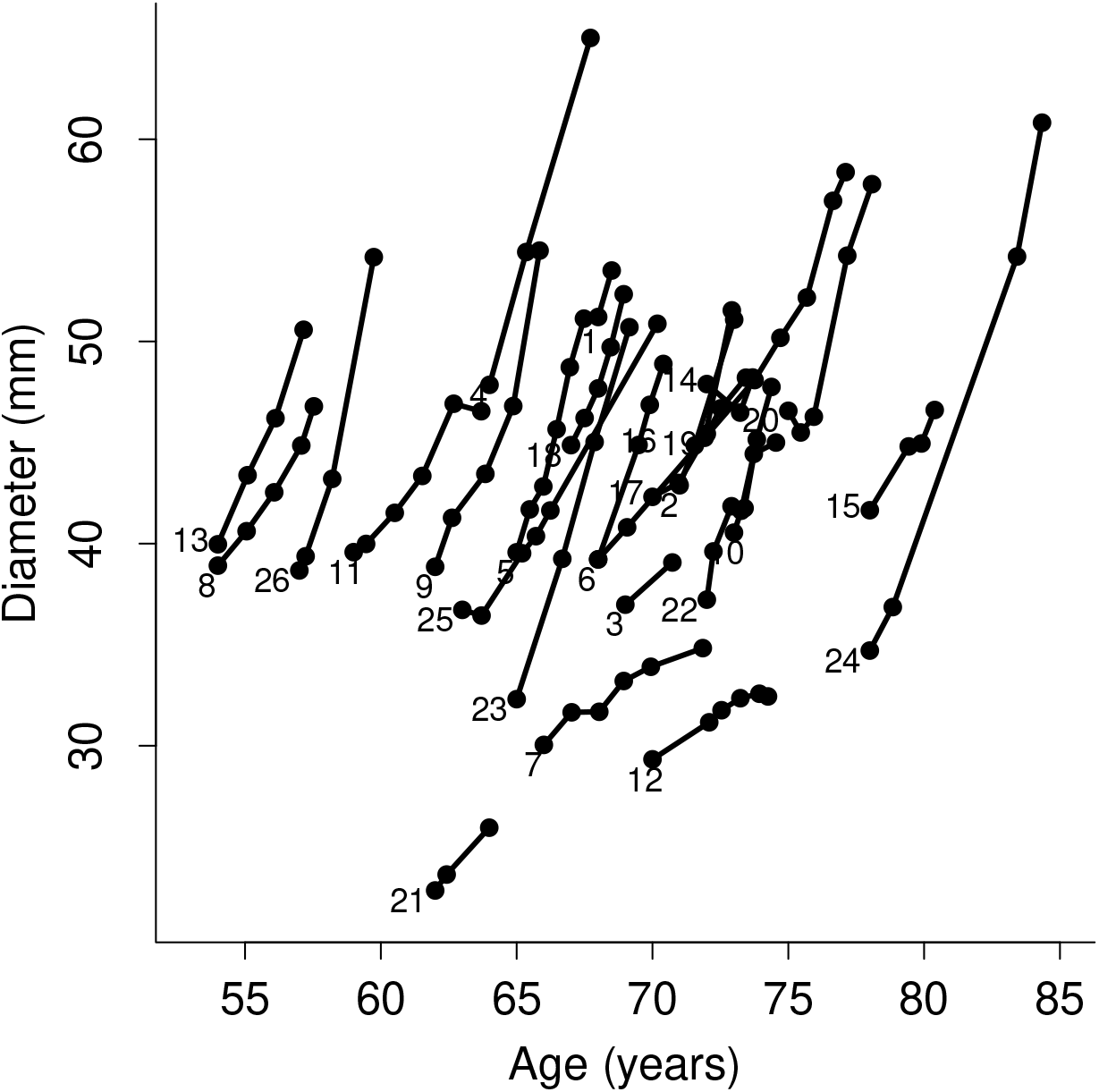
Plot of AAA spherical diameter vs. age for each patient. Each line corresponds to a patient. Patient ID is provided on the lower left corner of each line. Dots on each line indicate when the CT image was taken.

A multistate continuous-time Markov chain model is used to model the progression of diseases (see, e.g. Jackson *et al*., 2003^19^) which can provide information of disease progression by transition probabilities between different states and mean sojourn time at one state. Specifically, we used a homogeneous model by assuming a transition intensity (the rate of a transition probability) is independent of time. This model allows a transition probability that changes over time but the rate remains constant. In addition, we incorporate ILT information into the model by considering the logarithm of an intensity as a linear function of explanatory variables from ILT. To investigate which model fits the data better, we consider likelihood ratio (LR) statistics given the proposed statistical model. The LR statistics is the ratio of two likelihoods (or -2 log likelihood ratio), which is used to perform a statistical hypothesis test for model comparison. The likelihood in the numerator is from the null model and the likelihood in the denominator is from the alternative model. Thus, a larger value of the likelihood ratio statistics (or a smaller value of -2 log likelihood ratio) indicates the data support the null model. The estimation algorithm is converged produced the stable results.

## Results

Recall that we consider the maximal thickness of ILT (max_ILT_), the fraction of wall area covered by ILT (area_frac_) and the volume fraction of ILT (vol_frac_) into the model as covariates. These variables are continuous variables. Note that these three variables are highly correlated with each other. In particular, the correlation between the fraction of wall area covered by ILT and the volume fraction of ILT is 0.89. High correlation between covariates can lead unstable estimates if they are included in the model together. Thus, we consider a model with one covariate each. Although three variables are highly correlated, as shown in Figure 2, they have different effects on the transition probability curves at different times. We want to investigate which variable is a significant biomarker for the progression of AAA.

**Figure 2.**
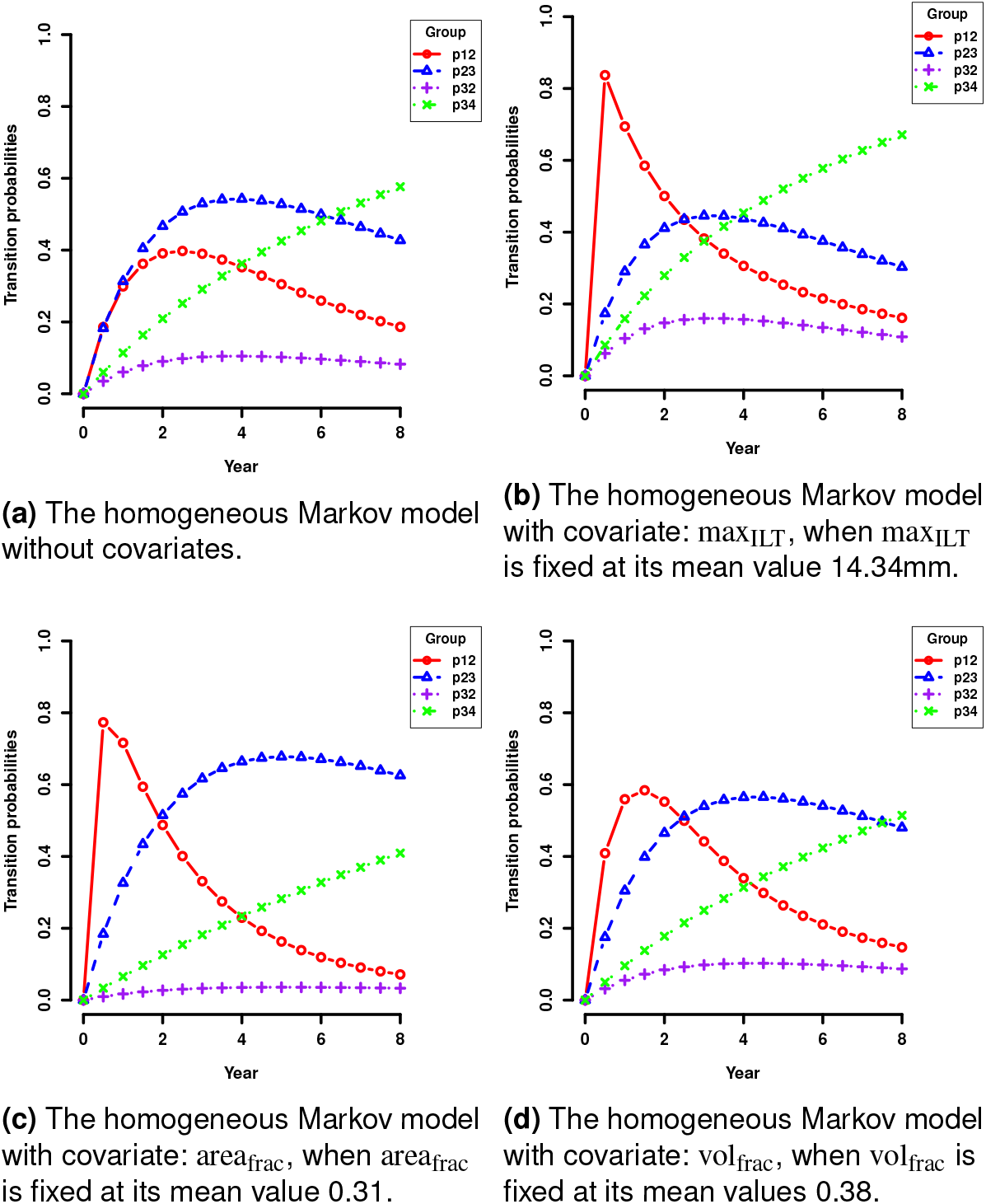
Fitted transition probability curves at different years. In each graph, the red circle line (p12) denotes transition probabilities of moving from ‘early’ state to ‘mild’ state; the blue triangle line (p23) denotes transition probabilities of moving from ‘mild’ state to ‘severe’ state; the purple plus line (p32) denotes transition probabilities of moving from ‘severe’ state to ‘mild’ state: the green cross line (p34) denotes transition probabilities of moving from ‘severe’ state to ‘fatal’ state. The x-axis is years. The y-axis is the transition probability.

### Model comparison

Table 1 shows likelihood ratio statistics (−2 log likelihood ratio) and the corresponding p-value for each model when the null model is the model without covariates. This result shows that the model with one of such covariates is overall statistically significant. Although we cannot directly compare which covariate fits the data better than the others by a hypothesis test as they are not nested models, the larger LR statistics could imply a better fit as the LR statistics of each model is calculated with the same null model.

**Table 1.** Likelihood ratio statistics of different models.

| | $\max_{\text{ILT}}$ | $\text{area}_{\text{frac}}$ | $\text{vol}_{\text{frac}}$ |
| --- | --- | --- | --- |
| -2 log LR | 17.9 | 23.33 | 10.51 |
| p value | 0.0013 | 0.0001 | 0.0326 |

Another evidence that these three covariates contribute much to the progress of the AAA enlargement is the comparison of transition probability curves under different models (see Figures 2a, 2b, 2c and 2d). The fitted transition probability curve without covariates has relatively lower peaks than all the others, especially when the transition is from the ‘early’ state to the ’mild’ state. In addition, by finding the peak of the curves *p*_12_ and *p*_23_, we conclude that patients in ‘early’ state are most likely to move to ‘mild’ state between 1 and 2 years, while patients in ‘mild’ state are most likely to move to ‘severe’ state around 4 years.

The model with area_frac_ has the highest transition probability from the ‘mild’ state to the ‘severe’ state among the three models given the severe criterion within 3 years (Figures 2b, 2c and 2d). This may indicate the progression from the ‘mild’ state to the ‘severe’ state is sensitive to the changes of area_frac_. In other words, when a patient already has a mild AAA, area_frac_ can be a useful biomarker to predict his/her state after a certain time.

A prevalence plot could also show goodness of fit for a multistate continuous Markov Chain model and Pearson tests can provide goodness of fit for the models as well. These results indicate that the Markov model with max_ILT_ or area_frac_ as a covariate passed the test of a goodness of fit. We provide detailed results in the supplementary document.

Among the models with ILT characteristics as a covariate, we investigate the model using wall area covered by ILT (area_frac_) with more details. Results of the other two models (max_ILT_ and vol_frac_) are provided in the supplementary document.

### Fraction of wall area covered by ILT

The value of area_frac_ ranges from 0 to 0.74, and about 75% of cases are less than 0.4. So the value 0.4 (75% quantile) could be used as a criterion for the severity of ILT. As an extreme case, we also consider 0.6 which is 95% quantile. With these values for area_frac_, we provide estimated transition probabilities for the duration of 3 years (Table 2a, 2b). Also we provide mean sojourn time in Table 2c. According to the estimated transition probability ((1,1)-th entry of Table 2a), we can see that there is no chance that a patient with area_frac_ being 0.4 is in ‘early’ state. Besides, such a patient will instantly transit to or already stay in either ‘mild’ or ‘severe’ state within 3 years. By comparing transition probabilities when area_frac_ is 0.6 (Table 2b) with those when area_frac_ is 0.4 (Table 2a), we also conclude that disease state of AAA progresses significantly with the increase of area_frac_. Also, mean sojourn time for ‘early’ state is significantly small compared to those for other states. Although the range of AAA size for each state is rather different, this still implies that a patient stays ‘early’ for a shorter time with 0.4 of area_frac_.

**Table 2.**
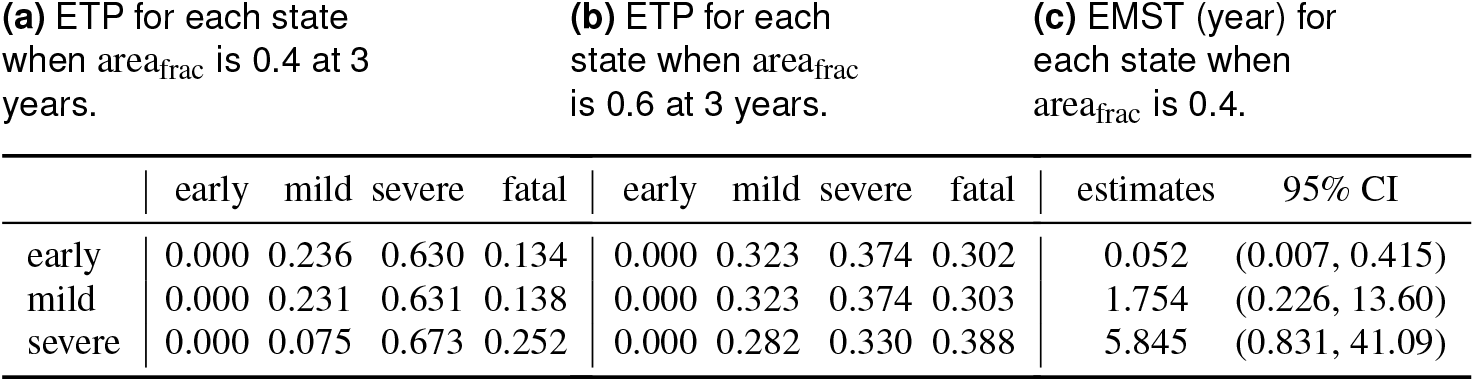
Results from multistate continuous-time Markov chain models with the covariate areafrac. (a) and (b) denote estimated transition probabilities (ETP) at the different values of the covariate and time. (c) denote the estimated mean sojourn time (EMST).

| (a) ETP for each state<br>when $\text{area}_{\text{frac}}$ is 0.4 at 3<br>years. | | | | | (b) ETP for each<br>state when $\text{area}_{\text{frac}}$<br>is 0.6 at 3 years. | | | | | (c) EMST (year) for<br>each state when<br>$\text{area}_{\text{frac}}$ is 0.4. | |
| --- | --- | --- | --- | --- | --- | --- | --- | --- | --- | --- | --- |
|  | early | mild | severe | fatal | early | mild | severe | fatal |  | estimates | 95% CI |
| early | 0.000 | 0.236 | 0.630 | 0.134 | 0.000 | 0.323 | 0.374 | 0.302 |  | 0.052 | (0.007, 0.415) |
| mild | 0.000 | 0.231 | 0.631 | 0.138 | 0.000 | 0.323 | 0.374 | 0.303 |  | 1.754 | (0.226, 13.60) |
| severe | 0.000 | 0.075 | 0.673 | 0.252 | 0.000 | 0.282 | 0.330 | 0.388 |  | 5.845 | (0.831, 41.09) |

To see the effects of area_frac_ on the survival probability, we plot fitted survival probability curves with Kaplan-Meier (KM) curves, the empirical estimate of the survival probability^20^. By comparing Figure 3a with Figure 3b, we find that the model with larger area_frac_ as a covariate has a lower survival probability which implies that a patient with larger area_frac_ is exposed to a higher risk of entering into the last stage (‘fatal’ state). This result is consistent with what we have found earlier. One can compare two survival curves, for example, the one with area_frac_ = 0.4 and the other with area_frac_ = 0.6 by two sample Kolmogorov-Smirnov test^21^, which indicates the two curves are statistically different. The detailed results of the test are provided in the supplementary document.

**Figure 3.**
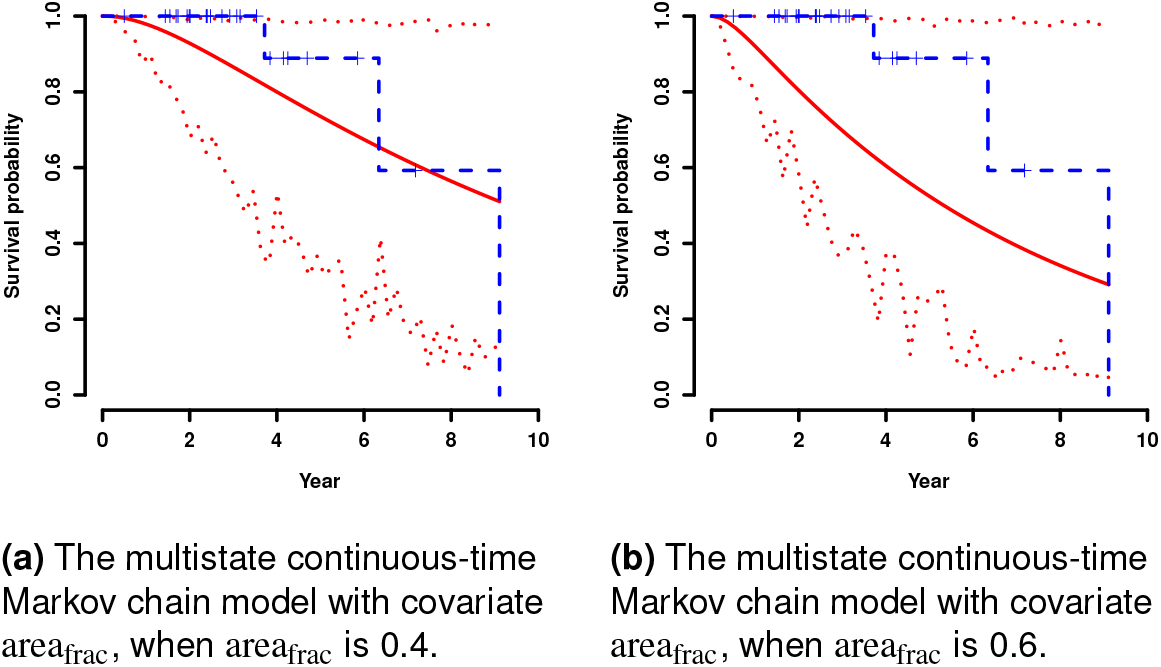
Comparison of empirical and fitted survival probability for two multistate continuous-time Markov chain models with areafrac. The red solid line denotes the fitted survival curve. The blue dashed line denotes the empirical survival. The red dotted line denotes the 95% confidence interval of the fitted survival.

A bigger set of patient data should be tested in order to reach a stronger conclusion. However, despite the fact that a small data set was used, a trend was clearly seen. If we believe that a surgical intervention should be recommended when survival probability lower than 40% at 4 years, Figure 3b suggests that a patient with 0.6 of areafrac is being considered for such recommendation. For maxILT, 30mm was the value we obtain for such recommendation. Note that depending on the practitioner’s criterion on the survival probability (e.g. 40%) and the time period (e.g. 4 years), the recommendation will change. However, the model we used can provide such information adaptively.

## Discussion

A homogeneous multistate continuous-time Markov chain model is considered for the analysis of AAA progression to investigate transition of progression from one state to the another state in AAA growth. The model allows to estimate mean sojourn time for a patient being in each state. Given the estimated mean sojourn times for ’mild’ and ’severe’ states from the model with area_frac_, we may say that an approximated time for 1mm increase in the ’mild’ state is shorter than that for the ’severe’ state, which implies that AAA expands relatively fast at an early stage but the rate gets slower once AAA is enlarged enough. The model with ILT area fraction has the highest transition probabilities among the three models within 3 years at the value of the third quartile (severe condition). This finding may imply that the progression from ‘mild’ state to ‘severe’ state is sensitive to the changes of ILT area fraction. In other words, when someone already has a mild AAA, ILT area fraction may be a useful biomarker to predict his/her state after a certain time.

Likelihood ratio statistics supports that the model with one of three covariates fits the data better than the model without ILT information. This may suggest that when we consider AAA growth, we should not ignore ILT information such as maximal ILT thickness, ILT area fraction, ILT volume fraction information of the patients. More specifically, if we believe that the patient with the fitted survival probability lower than 40% at 4 year should be recommended for surgical intervention, we may suggest any patients with ILT area fraction more than 60% is highly recommended for surgical intervention, considering the low survival probabilities in these two progression stages. This recommendation could be changed depending on the threshold of survival probability and the time. Also, the results would be different than the reported due to the dependency of the chosen stages threshold criteria. More detailed analysis on the appropriate threshold should be conducted with a larger cohort before reaching stronger conclusion. Despite this limitation, our results are in agreement with other previous studies such as Domonkos *et al*. (2019)^22^ and Metaxa *et al*. (2015)^23^.

The thrombus accumulation is an ongoing process presented in 75% of detected AAA and the prevalence of finding such accumulation increases as the aneurysm size increases^24^. It is, therefore, expected to observe AAAs without ILT at the early stage (early) of this disease. This study found that most of our AAAs were remained without ILT accumulation during the early stage and while they transitioned to the following stage, a significant ILT accumulation was observed. Over this early stage, a faster AAA expansion in comparison with other stages was found. This could suggest that the faster growth may promote ILT accumulation. Additionally, this observation and the fact that ILT accumulation rate was found to be the same to the AAA expansion rate^16^ would explain the proposed idea of using sudden increase of thrombus as a potential predictor of rupture.

Once the initial ILT accumulation occurs (usually in the transition between ‘early’ to ‘mild’ state), our results have shown that the next transition stage would be sensitive to the amount of lumen area covered by the thrombus (area_frac_). The intraluminal thrombus is a complex biological entity composed also by many inflammatory cells, including macrophages and neutrophils^25^ that not just interfere with the direct wall shear stress and strain relationship between lumen wall and blood flow but that also interact biochemically (promoting wall thinning, cell inflammation, degradation of the extracellular matrix^26^) and biomechanically (modifying wall shear stress^27^) with the arterial wall. It is then understandable that a larger area covered by thrombus would have a greater impact on the AAA growth process and thus in our transition time. Furthermore, our results may indicate that the risk of a patient to transition to the ‘fatal’ last stage is outweighed as a larger ILT thickness (max_ILT_) is observed. These results agree with signs of lower pO2 levels and signs of hypoxia in aneurysm with ILT thickness greater than 4 mm^28^.

The data were originally obtained for characterizing relevant morphological parameters so that the clinical and biochemical data such as smoking habit (current, ex-smoking and how long) and blood pressure were not diligently collected. Also, we have only three female patients so that we could not investigate the effect by gender. Thus, we were unable to add more patient-specific information. If we have enough observations from the control group (individual with no AAA) and patients whose AAA ruptured or received surgical treatment, we would have more reliable estimates of transition probabilities as well as mean sojourn times. Also, thorough investigation on the effects of ILT information as well as other patient-specific information to the AAA growth can be done with a larger size of data. Therefore, we would like to point out that our findings bring an interesting hypothesis which should be verified by further studies with a larger group of AAA patients.

On the other hand, to our knowledge, there were only a few studies, available for the morphological data, obtained from follow-up patients. In fact, this study was one of large data sets, in terms of characterizing morphological parameters in the follow-up study. The reason of the scarcity of such data should be due to requiring manual time consuming segmentation or development of automatic process of quantifying the morphological parameters. We think that, even with the small size of samples, we was able to demonstrate the new statistical tool in the role of thrombus effect and predicting the progression of AAAs. For a future study, we wish to get clinical data with various risk factors and we plan to develop automatic morphological quantification so that we can extract ILT information easily, which helps to make a larger data for the analysis.

A growth rate/expansion rate could be also an informative criterion to determine the stages of AAA as well. However, we decided to select a maximum diameter given that it is the clinically important metric to assess the onset, progression, and risk of rupture in AAAs. The use of this widely known relationship to stratify AAAs severity would make it easier for the reader to relate our results to other patients’ cohort or to patients in clinical practice.

## Data Availability

The data that support the findings of this study are available on request from the corresponding author.

## Acknowledgements

All the codes are written and performed in R (r-project.org version 3.4.4.). The R package that we mainly used is “msm” and usage is introduced in Jackson *et al*. (2011) ^29^.

